# Genetic ancestry contributes to somatic mutations in lung cancers from admixed Latin American populations

**DOI:** 10.1101/2020.09.01.20183913

**Authors:** Jian Carrot-Zhang, Giovanny Soca-Chafre, Nick Patterson, Aaron R. Thorner, Anwesha Nag, Jacqueline Watson, Giulio Genovese, July Rodriguez, Maya K. Gelbard, Luis Corrales-Rodriguez, Yoichiro Mitsuishi, Gavin Ha, Joshua D. Campbell, Geoffrey R. Oxnard, Oscar Arrieta, Andres F. Cardona, Alexander Gusev, Matthew Meyerson

**Affiliations:** Department of Medical Oncology, Dana-Farber Cancer Institute, Boston, MA, USA; Broad Institute of MIT and Harvard, Cambridge, MA, USA; Departments of Genetics and Medicine, Harvard Medical School, Boston, MA, USA; Personalized Medicine Laboratory, Instituto Nacional de Cancerología, México City, México; Foundation for Clinical and Applied Cancer Research - FICMAC, Bogotá, Colombia; Medical Oncology, Hospital San Juan de Dios, San José, Costa Rica; Centro de Investigación y Manejo del Cáncer - CIMCA, San José, Costa Rica; Division of Respiratory Medicine, Graduate School of Medicine, Juntendo University, Bunkyo-ku, Tokyo, Japan; Division of Public Health Sciences, Fred Hutchinson Cancer Research Center, Seattle, WA, USA; Division of Computational Biomedicine, Department of Medicine, Boston University School of Medicine, Boston, MA, USA; Thoracic Oncology Unit, Instituto Nacional de Cancerologia, México City, México; Clinical and Translational Oncology Group, Clínica del Country, Bogotá, Colombia; Division of Genetics, Brigham and Women’s Hospital, Boston, MA 02115, USA

## Abstract

Inherited lung cancer risk, particularly in non-smokers, is poorly understood. Genomic and ancestry analysis of 1,153 lung cancers from Latin America revealed striking associations between Native American ancestry and their somatic landscape, including tumor mutational burden (TMB), and specific driver mutations in *EGFR, KRAS*, and *STK11*. A local Native American ancestry risk score predicted *EGFR* and *KRAS* mutation frequency more strongly than global ancestry, suggesting that germline genetics (rather than environmental exposure) underlie these disparities.

**Significance:** The frequency of somatic *EGFR* and *KRAS* mutations in lung cancer varies by ethnicity but we do not understand why. Our study suggests that the variation in *EGFR* and *KRAS* is directly associated with genetic ancestry and suggests further studies to identify germline alleles that underpin this association.

## Introduction

Lung cancer causes over 1.7 million deaths per year world-wide^1^, and kills more people than any other malignancy in Latin America^2^. Lung adenocarcinoma (LUAD) is the most common subtype of lung cancer that is typically driven by genomic alterations of genes in the receptor tyrosine kinase (RTK)/RAS/RAF pathway^3^ often allowing effective therapeutic targeting by RTK and other pathway inhibitors. It is well-known, but mysterious, that the frequency of somatic *EGFR* mutations is higher in LUADs from patients in East Asia (~45%) compared to LUADs from patients in Europe or patients of European (EUR) and/or African (AFR) descent in North America (~10%)^4-8^. In Latin American countries, the frequency of somatic *EGFR* mutations in LUAD ranges from roughly 14% in Argentina, to 25-34% in Colombia, Brazil and Mexico, to 51% in Peru^9-11^ (**Fig. 1A**). Moreover, recent genomic studies from East Asian (EAS) and African (AFR) populations have suggested different distributions of tumor mutation burden (TMB) and levels of somatic copy number alteration (SCNA)^12,13^, compared to LUAD patients of European (EUR) ancestry.

**Fig. 1:**
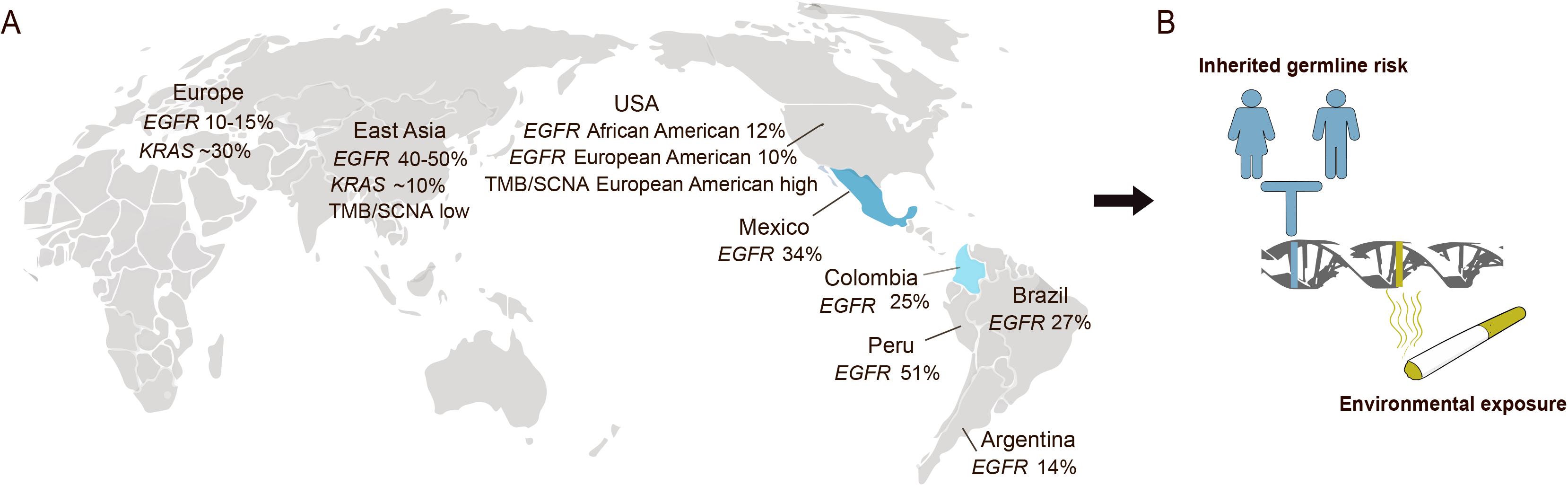
Genomic differences in LUAD across patient populations. A) In lung cancers from East Asian patients, TMB, SCNA burden and the frequency of *KRAS* mutations are lower, while the frequency of *EGFR* mutations is higher, compared to somatic genome alterations in lung cancers from patients of European and/or African origin. The somatic *EGFR* mutation rate in lung cancer varies among Latin American countries. B) Both germline variations and environmental exposures such as smoking can predispose to somatic alterations driving the development of lung cancers that may cause the genomic differences across populations.

Despite the differences in patterns of somatic mutation between LUAD from patients of different ethnicity, the landscape of ancestry effects on the lung cancer genomes for the Latin American populations has not been comprehensively described; and it remains unknown whether the differences are due to ancestry-specific germline variation, or rather to population-specific environmental exposures (**Fig. 1B**). This is of particular importance as Native American (NAT) ancestry -- which includes components of East Asian (EAS) ancestry derived through waves of migration^14^ - is present to varying degrees in modern populations in Latin America, along with EUR and AFR ancestry^15^.

## Results

To explore the landscape of somatic cancer mutation in lung cancers from Latin America and to assess the influence of germline ancestry of genetically admixed patient populations on these somatic alterations, we performed genomic analysis of 601 lung cancer cases from Mexico and 552 from Colombia, including 499 self-reported non-smokers. Next-generation sequencing targeting a panel of 547 cancer genes plus intronic regions of 60 cancer genes^16^ was used to identify single nucleotide variants (SNVs), indels, SCNAs) and gene fusions. This gene panel covers all currently known lung cancer drivers^3^, which are the focus of this work. For 896/1153 (78%) samples, we achieved at least 30X coverage in 80% targeted regions. We applied a custom script to identify hotspot driver mutations (**Methods**), and found that 538 (46%) of all samples harbored oncogenic mutations in *EGFR, KRAS, BRAF, ERBB2*, or *MET*, or fusions in *ALK*(**Fig. 2**). SCNA analysis (**Methods**) identified 9% and 2% cases with high-level amplifications in *MYC* and *MDM2*, respectively. We did not observe novel amplification or deletion peaks in this Latin American lung cancer cohort as assessed by GISTIC analysis.

**Fig 2:**
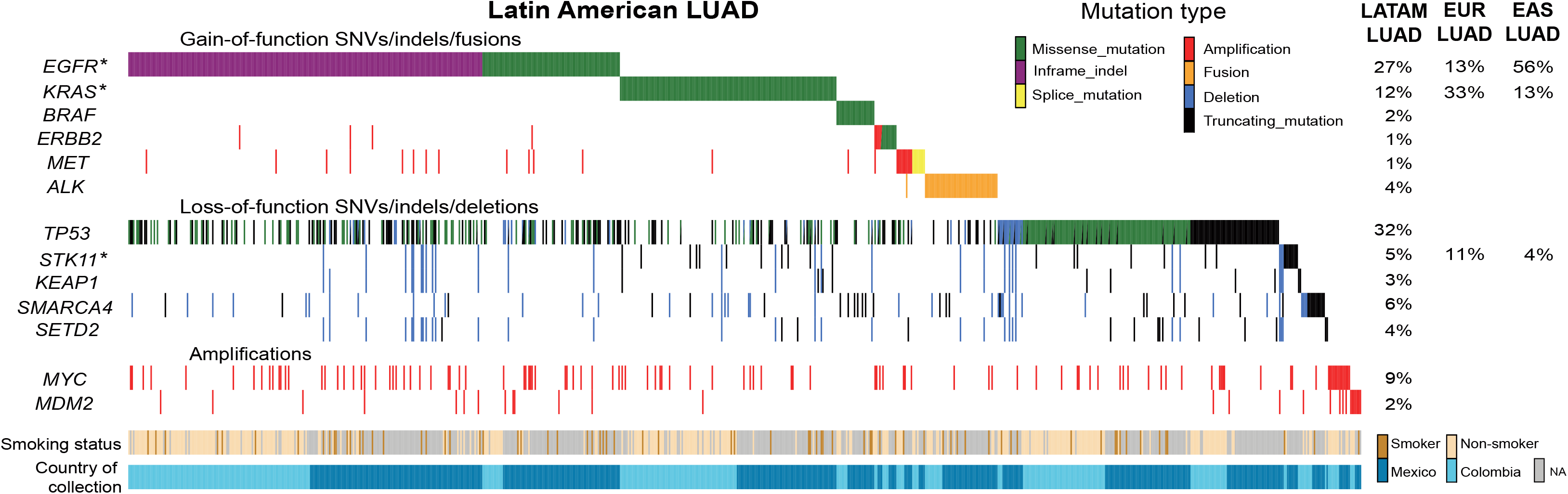
Somatic genome analysis of lung cancers from Mexico and Colombia. Co-mutation plot displays for known activators of the RTK/RAS/RAF pathway, tumor suppressor genes and significantly amplified genes. * indicates that oncogenic mutations in *EGFR* and *KRAS* as well as truncating mutations in *STK11* are associated with NAT ancestry, but other somatic alterations are not; correlations with mutations are controlled for TMB. LATAM: Latin American. The mutation frequency for EUR LUAD is obtained from the TCGA dataset. The mutation frequency for EAS LUAD is obtained from Chen et al 2020.

Ancestry effects on somatic cancer genomes are understudied, largely because both germline and somatic data from the same patients have generally been required^17,18^. We developed an analytical pipeline (https://github.com/jcarrotzhang/ancestry-from-panel) that offers the advantage of simultaneous measurement of global and local ancestry from sequencing tumor DNA only (**Fig. S1**). Briefly, we called the genotype of germline single nucleotide polymorphisms (SNP) using on-target and off-target reads from the sequencing panel, and measured global ancestry based on principal component analysis (PCA)^19^ of the germline SNPs, in which principal components (PCs) 1, 2 and 3 captured prominently the axis of AFR, EUR and NAT ancestry, respectively (**Fig. S2A**). We then imputed missing SNPs using an external haplotype reference panel^15^, and assigned local ancestry to each genomic region^20^, based on the imputed variants. We validated our approach to ancestry analysis by comparing tumor and normal ancestry estimations (Pearson’s r>0.99), and by comparing panel sequencing to whole-genome sequencing (Pearson’s r>0.96, **Fig. S2B-C**).

Having obtained data on both somatic alteration and genetic ancestry, our next step was to assess the correlation of these features, using multivariate regression controlling for self-reported smoking status and country of sample collection (Methods). As previous work focused on differences *between* populations, these associations with ancestry *within* a single admixed population provide more direct evidence of a putative genetic cause. First, we found a significant anti-correlation between TMB and PC3 representing the NAT ancestry (P=1×10^-6^, coef.=-0.02), in line with previous study of lung cancers from EAS patients^12^; no correlation was found with the total SCNA burden, or with aneuploidy.

Evaluation of ancestry-mutation association, adjusting for sample-specific TMB, in each gene from **Fig 2** showed that NAT ancestry was positively correlated with mutations in *EGFR* (FDR q=7×10^-5^, coef.=0.005), and anti-correlated with mutations in *KRAS* (FDR q=7×10^-5^, coef.=-0.007), and mutations in *STK11* (FDR q=5×10^-4^, coef.=-0.013). Each feature (TMB, *EGFR, KRAS*, or *STK11* mutation) was independently associated with NAT ancestry in a joint model (**Table S2**). Further analysis demonstrated that NAT ancestry was predominantly associated with oncogenic, driver mutations in *EGFR*, but not with non-oncogenic, passenger mutations (**Fig. S3**), suggesting an underlying mechanism influencing tumor fitness rather than mutagenesis. In addition, we did not observe SCNA of any lung cancer driver gene associated with ancestry (Methods). The observed correlations held in separate analyses of the Mexican and Colombian cohorts (**Fig. S4**).

To better understand the relationship of ancestry and exposure-induced mutagenesis in the risk of developing lung adenocarcinomas through the activation of RTK/RAS/RAF pathway, we tested the ancestry associations in RTK/RAS/RAF pathway oncogenes adjusting for mutational signatures (**Table S3**). The positive correlation of NAT ancestry with *EGFR* mutation (OR=1.23 in every 10% increase of NAT ancestry, 95% CI 1.12-1.35), and negative correlation of NAT ancestry with *KRAS* mutation (OR=0.85 in every 10% increase of NAT ancestry, 95% CI 0.77-0.95) remained significant (**Fig. 3A, 3B**). The association with *EGFR* held in an analysis restricted to patients who reported never smoking (OR=1.46 in every 10% increase of NAT ancestry, 95% CI 1.25-1.70, **Fig. 3B**). When including smokers, *KRAS* mutation rate increased with the proportion of smoking signature (OR=1.27 in every 10% increase of smoking signature, 95% CI 1.04-1.56, **Fig. 3B**). The ancestry effect on *KRAS* mutations in reported never smokers trended toward significance but was not significant (P=0.08) in this study, perhaps due to sample size. Moreover, the interaction of smoking signature and NAT ancestry did not modify the effect size of ancestry on *KRAS* (Methods). Gender and the APOBEC signature were not associated with mutations of any lung cancer oncogenes. Age of diagnosis was negatively associated with the risk of ALK-translocated cases (P=3×10^-5^, 0R=0.97 in every 10-year increase of age, 95% CI 0.96-0.99). Together, we conclude that NAT ancestry was associated with genomic differences in Latin American LUAD patients that are independent of smoking activity.

**Fig. 3:**
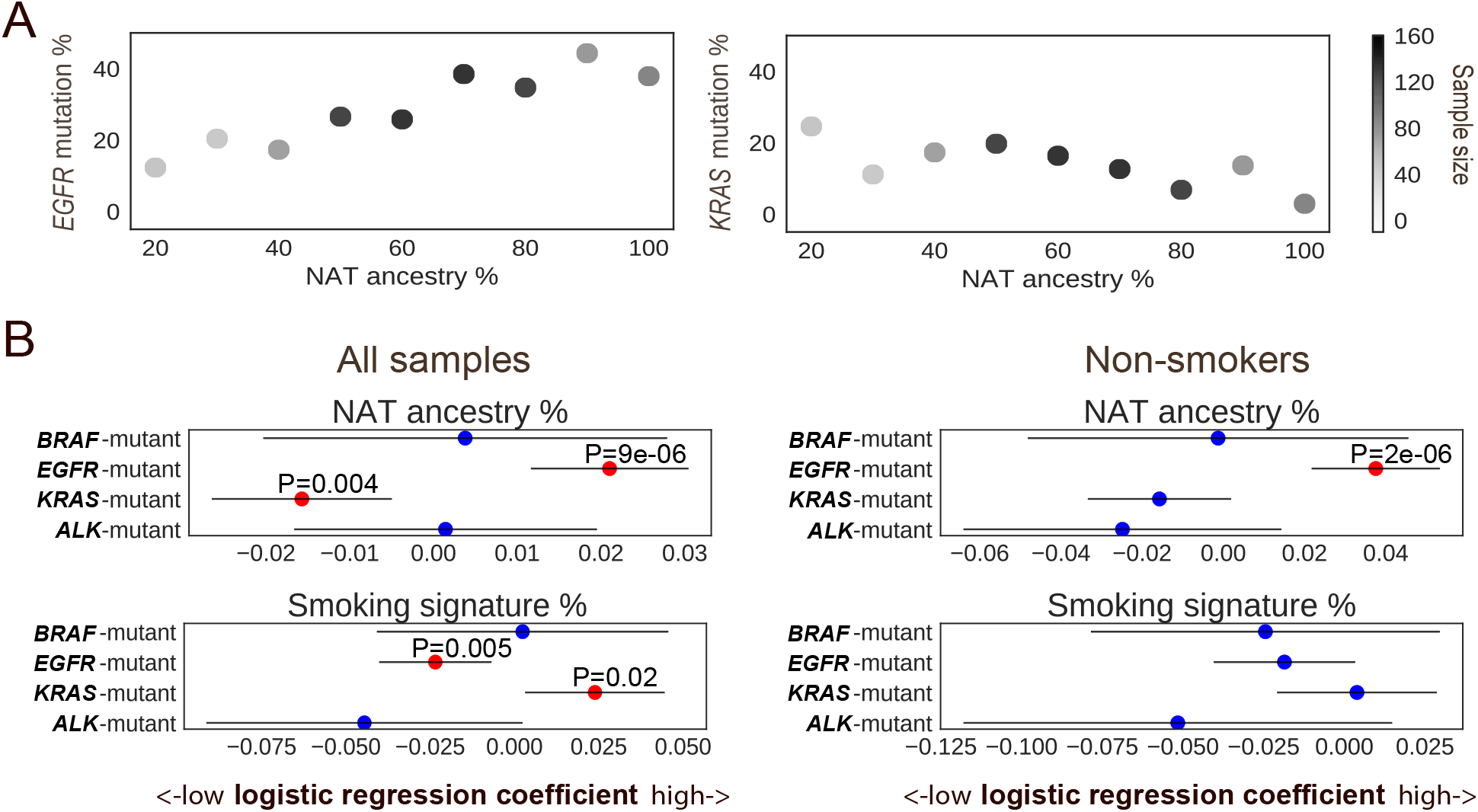
Targetable LUAD driver genes associated with genetic ancestry. A) The percentage of NAT germline ancestry is positively correlated with the percentage of somatic *EGFR* mutations, and negatively correlated with the percentage of somatic *KRAS* mutations. Color bar represents the number of samples in the NAT ancestry percentage range. B) Association of targetable LUAD driver genes with ancestry, mutational signature and gender in all cases (n=705) (left) and in never smokers only (n=387) (right). Multivariate logistic regression P values are shown, with NAT ancestry percentage, gender, smoking and APOBEC signature as covariates. Red dots represent FDR corrected q<0.1. Lines represent 95% confidence interval.

To assess whether the observed association with *EGFR* and *KRAS* mutations is due to NAT ancestry itself or to an environmental exposure/socioeconomic status related to the NAT ancestry, we next investigated the influence of local ancestry. Previous work has shown that associations between local ancestry and phenotype (while accounting for global ancestry) provide evidence of a genetically driven phenotype, as local ancestry is not expected to be non-causally associated with environmental exposure or socioeconomic status^21,22^. We used RFMix to map local ancestry, producing 5,059 genomic regions with an assignment of AFR, EUR or NAT ancestral population for each parental chromosome (**Table S4**). We performed a multivariable logistic regression of NAT ancestry for each genomic region correlating with the EGFR-mutant or KRAS-mutant samples, controlling for the global ancestry (Methods). We did not identify any region where correlation reached genome-wide significance of P<1×10^-5^ (**Fig. 4A, S5**).

**Fig 4:**
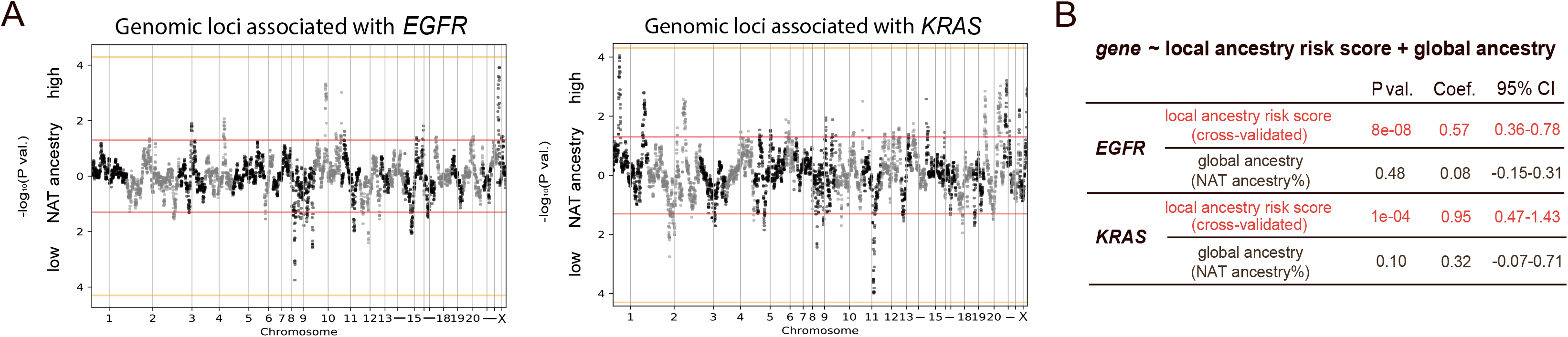
Germline local ancestry in association with somatic *EGFR* and *KRAS* mutations. A) Genome-wide association of local NAT ancestry with *EGFR* (left) and *KRAS* (right). “NAT ancestry high” indicates positive association, whereas “NAT ancestry low” indicates negative association. Red line indicates P=0.05. Orange line indicates genome-wide significance threshold (P<5×10^-5^). B) Association of local ancestry risk score with somatic *EGFR* or *KRAS* mutations, controlling for global ancestry (proportion of overall ancestry).

We next evaluated whether local ancestries across multiple sub genome-wide significance threshold regions (1×10^-5^<P<0.05) were associated with the somatic mutation phenotype by constructing a polygenic ancestry score. This approach is conceptually similar to previous work leveraging local ancestry to quantifying phenotypic heritability^21^, but we employ a risk score rather than variance partitioning as the former is more stable at low sample sizes. The local ancestry risk score was defined as the sum of NAT ancestry across each associated region weighted by the Z-score for the association of that region with the given mutation (Methods). To guard against overfitting, Z-scores and ancestry scores were computed by cross-validation: splitting the dataset into ten subsets, obtaining Z-scores for the mutation-ancestry associations using nine subsets, and then calculating the local ancestry risk score on the held-out subset (**Fig. S6**). We then performed another logistic regression including both the cross-validated local ancestry risk score and global ancestry as covariates, and found that the local ancestry risk score was significantly associated with *EGFR* and *KRAS* mutations, respectively, whereas global ancestry was no longer significant in the joint model (**Fig. 4B**). In contrast, the local ancestry risk score was not associated with TMB and *STK11* mutations in a joint model with global ancestry. Our finding suggests that multiple genetic loci specific to NAT ancestry may modulate the evolution of lung cancer tumors to harbor *EGFR* or *KRAS* mutations in the Latin American populations.

## Discussion

In summary, the genomic landscape of LUADs is strikingly varied in Latin American patients with mixed ancestries. In our study of 1,153 lung cancers, we demonstrated that NAT ancestry was correlated with somatic driver alterations, including *EGFR* and *KRAS* mutations that can be effectively targeted by small molecule inhibitors to prolong survival^4,23^, and TMB and *STK11* that are potential prognostic biomarkers in lung cancer patients^24,25^. The ancestry and TMB association were independent of smoking-related mutational processes, and therefore, further investigation on the impact of ancestry-related TMB differences on the response to checkpoint inhibitors is needed^26^. Of note, our TMB estimates may be susceptible to germline contaminations due to the lack of matched normal samples. If germline variants specific to the Mexican or Colombian population could not be sufficiently filtered (due to smaller germline reference panels), individuals with higher NAT ancestry would have more germline contamination and the anti-correlation between TMB and NAT ancestry may thus be even more significant than we have observed.

Our study provides the first example, to our knowledge, of a germline influence on targetable somatic events in lung cancer. As low-dose CT scans have enabled lung cancer screening that can significantly reduce lung cancer mortality^27,28^, the identification of germline ancestry predisposing to the development of EGFR-mutant or KRAS-mutant LUAD may shed light on the prevention and early detection strategies for admixed lung patients, particularly for non-smokers. Given the limited sample size, we could not determine the precise risk loci for *EGFR* and *KRAS* by local ancestry mapping. We believe that association analysis from larger lung cancer sample sets will have the power to identify a germline risk allele specific to the distinct cancer phenotype.

## Data Availability

Source data is provided as Supplementary Tables. Raw sequencing data is available for research purpose upon request.

## Acknowledgements

We would like to acknowledge the patients from Mexico and Colombia for their participation in this study. This study is supported by the V Foundation Translational Research Award and by National Cancer Institute grants R35 CA197568 and R01 CA116020. M.M. is an American Cancer Society Research Professor. J.C.-Z. holds the Banting fellowship. J.D.C. is funded by the LUNGevity Career Development award. We thank the Center for Cancer Genome Discovery at Dana-Farber Cancer Institute and the Genomic Platform at Broad Institute for their sequencing and genotyping efforts, and Aruna Ramachandran for the helpful discussions.

## Methods

### Sample collection

The protocol of this work was approved by the ethical and scientific committee in the Instituto Nacional de Cancerologia in Mexico City, the Foundation for Clinical and Applied Cancer Research in Bogotá, and Dana-Farber Cancer Institute in Boston for detecting *EGFR* mutations and further genomic analysis. Biopsies were collected for histological diagnosis by the pathology departments of Instituto Nacional de Cancerologia and Foundation for Clinical and Applied Cancer Research.

### Library preparation and sequencing

Genomic DNA was extracted from fresh-frozen, blood and paraffin-embedded samples by a standard procedure using the Wizard Genomics DNA kit (Promega, Madison, USA) according to the manufacturer’s instructions. DNA was fragmented to 250bp and size-selected DNA was ligated to sample-specific barcodes. A custom targeted hybrid capture sequencing platform (OncoPanel) was used to assay genomic alteration from tumor DNA^16^. Each library was quantified by sequencing on an Illumina MiSeq nano flow cell (Illumina, San Diego, CA). Libraries were pooled in equal mass to a total of 500ng for enrichment using the Agilent SureSelect hybrid capture kit (Agilent Technologies, Santa Clara, CA; cat. no. G9611A). Libraries were sequenced on an Illumina HiSeq2500 or HiSeq3000. Pooled samples were demultiplexed using the Picard tools (https://broadinstitute.github.io/picard/). Paired reads were aligned to the hg19 reference genome using BWA^29^ with the following parameters “-q 5 -l 32 -k 2 -o 1”. Aligned reads were sorted and duplicate-marked using Picard. In each batch, we sequenced a control DNA sample as a “plate normal”. For a subset of cases, the same libraries were sequenced on Illumina NovaSeq for low-coverage whole-genome sequencing.

### Mutation analysis

Mutation detection for single nucleotide variants (SNVs) was performed using MuTect v1.1.4^30^ in paired mode by pairing each sample to a control DNA sample profiled with the same OncoPanel. SomaticIndelDetector^31^ was used for indel calling. Mutations were annotated by Variant Effect Predictor (VEP) and Oncotator^33^. Called variants that were found in the Exome Aggregation Consortium (ExAC)^34^ with a frequency greater than or equal to 0.01% were excluded. TMB was calculated by dividing the total number of coding, non-silent mutations in an individual divided by the target size (3 MB). Mutational signatures were called using SignatureAnalyzer^35^ with SNVs classified by 96 tri-nucleotide contexts. Smoking and APOBEC signature activities are inferred by the estimated number of mutations in a tri-nucleotide context associated with each signature.

A custom script was applied to inspect the sites of hotspot driver mutations in *EGFR, KRAS, BRAF, ERBB2, MET*, and *TP53*. For each mutation, we counted reads supporting the reference base and the altered base, after filtering out reads with base quality or mapping quality less than 20. A mutation was called if the total read count was greater than 5, the altered read count was greater than 2, and the mutant allele frequency was greater than 5%. Identified mutations with total coverage lower than 30X were manually inspected using IGV^36^.

### Copy number and rearrangements

Read coverage was calculated at 1MB bins across the genome and was corrected for GC content and mappability biases using ichorCNA version 0.1.0^37^ using the plate normal as the matched control for each sample. GISTIC version 2.0.22^38^ was applied to identify focal and arm-level SCNAs on ichorCNA generated copy number segments, with the high-level amplification defined as log_2_-transformed copy number ratio greater than 0.7. Rearrangement events were called by Breakmer^39^ and filtered on discordant read counts and split read counts greater than 0. Total SCNA burden and the degree of aneuploidy was defined by the number of genes, or chromosomal arms affected by SCNAs, respectively (copy number ratio > 0.1 or copy number ratio > −0.1).

### Ancestry analysis from genotyping array

Multi-Ethnic Global Array (MEGA) was used for genotyping of paired fresh-frozen tumor tissue and blood samples. We used PLINK version 1.9^40^ to filter out variants with missingness greater than 2%, or failed Hardy-Weinberg equilibrium test (p<1e-06). Markers with allele frequency less than 1% in the 1000 Genomes dataset were also excluded. Our Mexican and Colombian samples were merged with samples from the 1000 Genomes phase 3^15^, and PCA was performed on the merged data set using (GCTA) version 1.91.6.^41^

### Ancestry analysis from sequencing

SAMtools^42^ was used to genotype germline variants after filtering out reads with base quality or mapping quality less than 20. LASER version 2.04^43^ was used to estimate overall ancestry based on 637,037 germline variants from all populations in HGDP^44^. We obtained principal components from LASER results that place each sample into a reference PCA space using 939 HGDP samples as reference samples. For local ancestry identification, phasing and imputation were performed using Beagle version 4.1^45^ based on SAMtools genotyped variants. We imputed missing variants using phased haplotypes from 1000 Genomes^15^. Ancestry was assigned to each SNP using RFMix v2^20^. For each parental population (NAT, AFR and EUR), 500 samples from 1000 Genomes were used as reference samples. Local ancestry regions spanning centromeres were filtered. RFMix outputted global ancestry estimates were used as the percentage of NAT ancestry.

### Association analysis

Multivariate logistic regression or linear regression was performed using a Python module (http://www.statsmodels.org). Because the Mexican population of lung cancer patients has higher level of NAT ancestry than the Colombian population^15^, we accounted for the country of sample collection throughout our analyses. TMB was included as a covariate when associating PC3 to mutations. Total SCNA burden was included as a covariate when associating PC3 to SCNA of lung cancer genes. Gender, proportion of smoking and APOBEC signatures were included as covariates when associating the percentage of NAT ancestry with oncogenic mutations. To test whether smoking signature influence the relationship between ancestry and the *KRAS* mutations, the following model was performed:

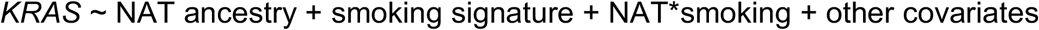

Where gender and country of sample collection were considered as covariates, and NAT*smoking was included as an interaction effect. If the interaction term is not significant, that means that smoking signature activity does not modify the effect of ancestry.

To identify specific genomic region(s) associated with LUAD cases harboring certain somatic alterations, a logistic regression model was applied controlling for the percentage of NAT ancestry, TMB and country of sample collection, followed by genomic control 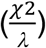. Ten-fold cross-validation was performed in the following steps: the whole dataset was split into ten subsets. Z-scores for the mutation-ancestry associations for each genomic region were calculated using nine subsets, and a cross-validated local ancestry risk score (sum of the NAT ancestry across each associated region weighted by the Z-score of that region) was calculated for each sample on the held-out subset. These steps were repeated for ten times until a local ancestry risk score was generated for each sample:

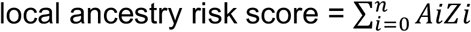

Where n is the number of regions associated with the somatic feature (P<0.05), A is the ancestry of each associated region (NAT ancestry was coded as 1, and EUR or AFR ancestry was coded as 0), and Z is the z-score of that associated region.

---

**Fig. S1:**
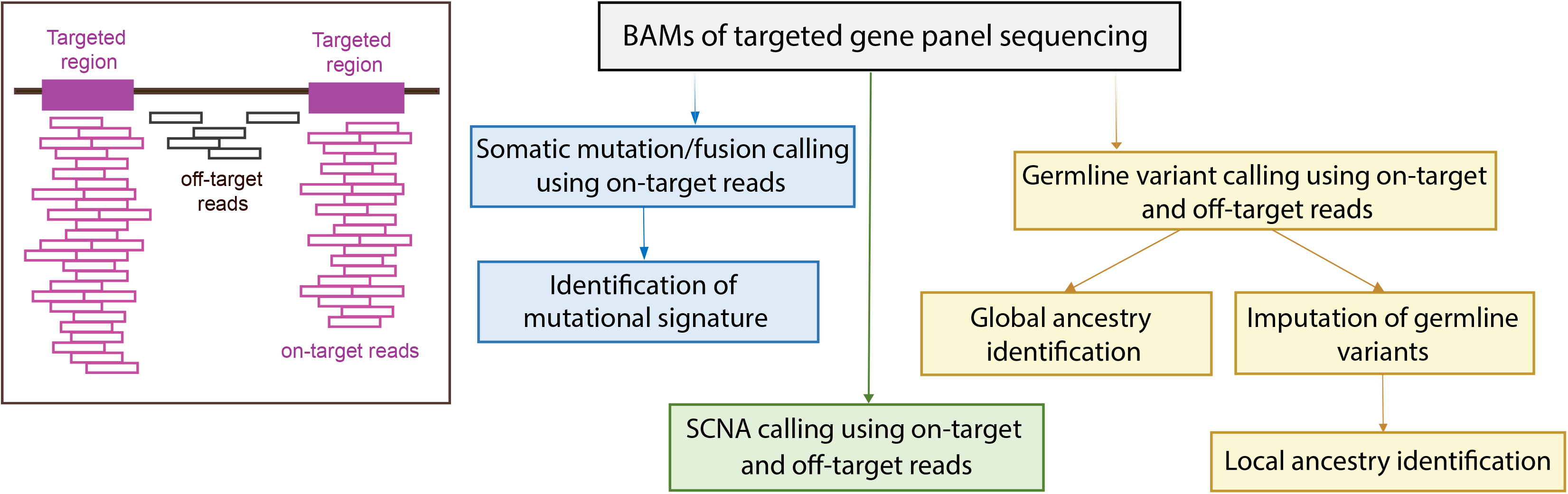
Overview of analytical pipeline. On-target and off-target reads are obtained from BAM files. On-target reads are used to identify somatic SNVs/indels/fusions. Both on-target and off-target reads are used to identify SCNAs, and to infer overall genetic ancestry (global ancestry) and germline SNP genotypes. Missing SNPs are imputed and then ancestry is assigned to each genomic region (local ancestry identification).

**Fig. S2:**
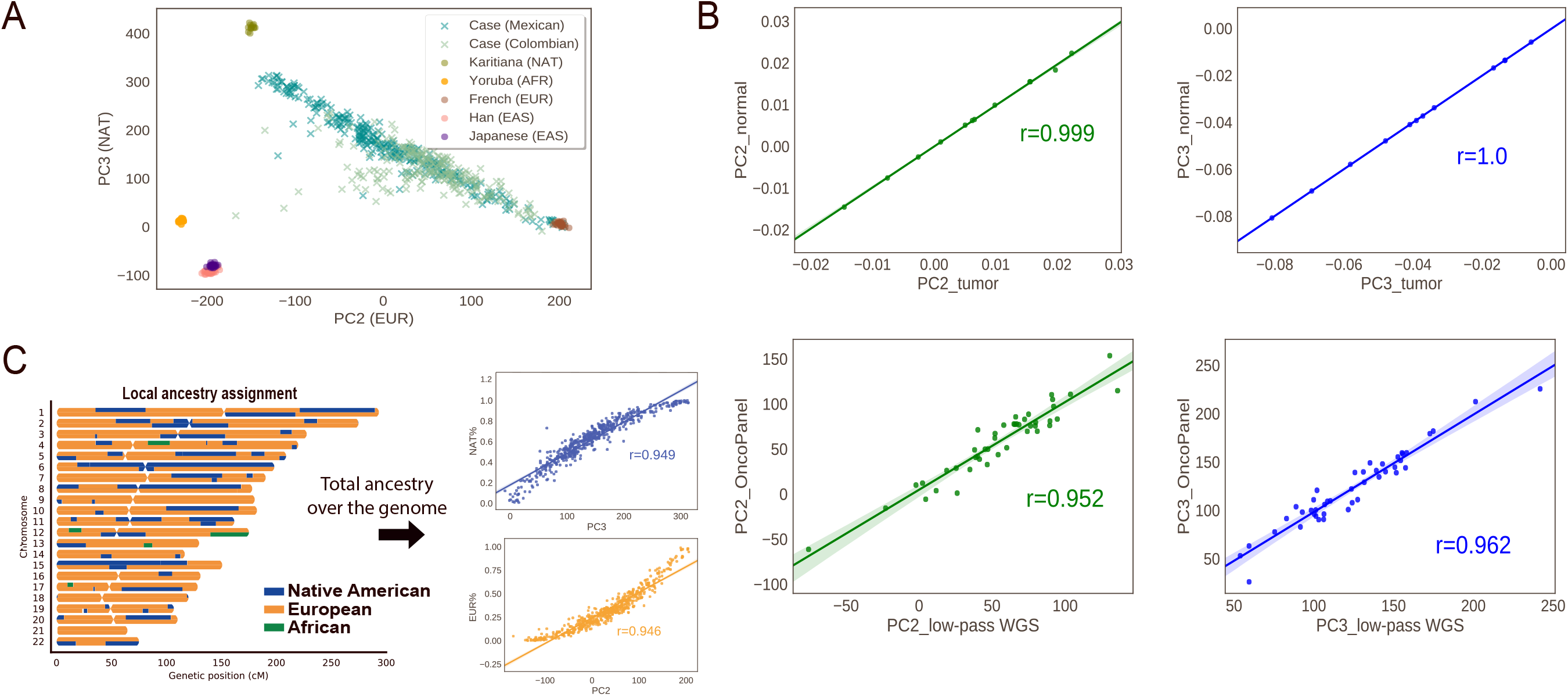
Validation of analytical pipeline. A) PC2 vs. PC3 from PCA of the Latin American cases in this study analyzed together with 939 reference samples from HGDP. The Latin American cases stretch out along PC2 and PC3, indicating the admixture of ancestries. B) High correlation of PC2 and PC3 between genotyping data of paired tumor vs. normal DNA from 12 patients (upper panel); and between the two sequencing approaches, panel vs. low-pass whole genome sequencing (lower panel) in 44 cases. C) An example of local ancestry identification. Each genomic region is assigned with NAT, EUR or AFR ancestry. The proportion of NAT and EUR ancestry per sample obtained from local ancestry highly correlates with PC3 and PC2, respectively. The r values are obtained from Spearman’s correlation.

**Fig. S3:**
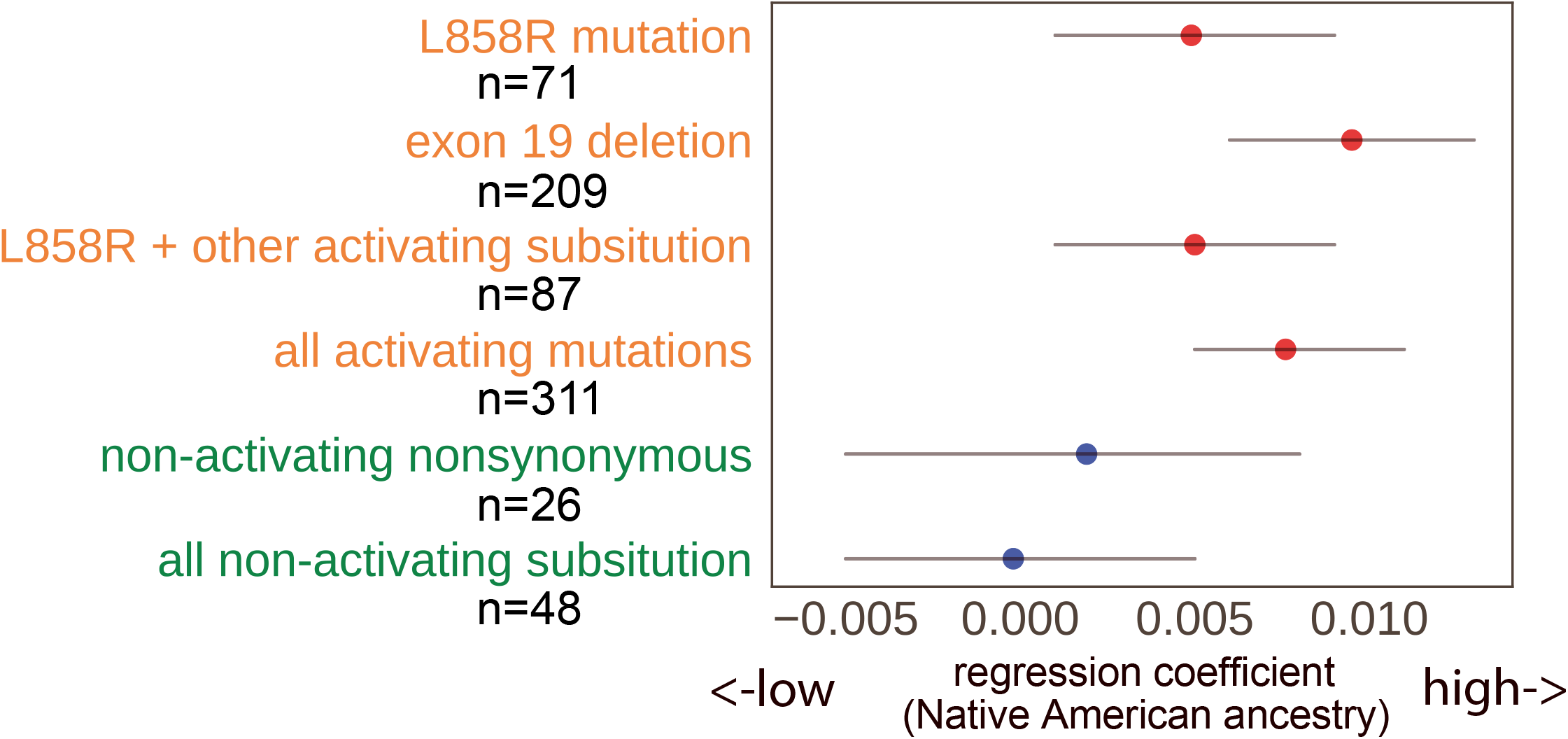
*EGFR* mutations in association with NAT ancestry. Logistic regression is used, coding EGFR-mutant cases as 1 and other oncogene-mutant cases as 0, and accounting for country of sample collection. Logistic regression coefficients are denoted by dots and 95% CIs are denoted by lines. Red dots represent correlations with P values less than 0.05. Sample size (n) for each mutation group of *EGFR* is indicated.

**Fig. S4:**
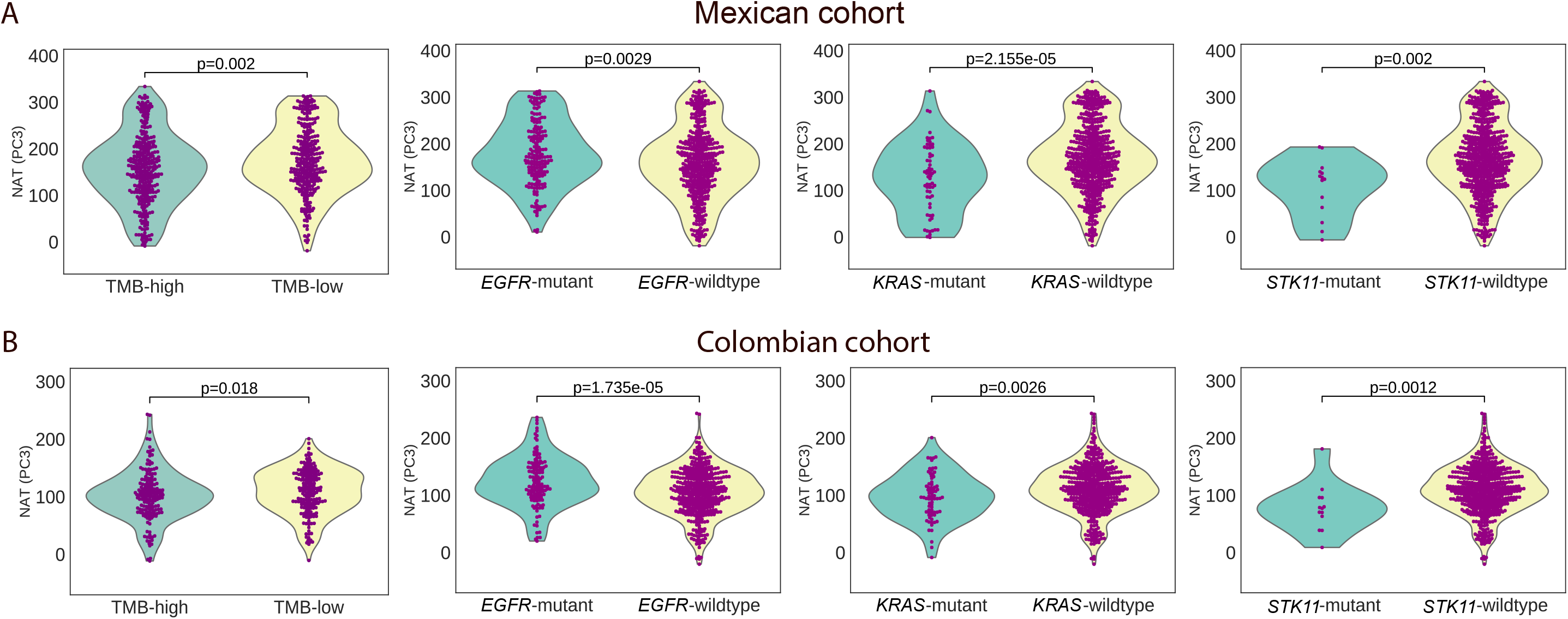
Correlation of genetic ancestry and somatic mutation in Mexican and Colombian samples separately. Comparison of PC3 indicating the NAT ancestry in cases with TMB-high and TMB low, and in cases with or without *EGFR, KRAS* or *STK11* mutations in A) the Mexican cohort and B) the Colombian cohort. TMB great than the median TMB for each cohort is defined as TMB-high. PC3 is obtained from PCA of all Mexican and Colombian cases with 939 HGDP samples as reference. P values are obtained from Mann-Whitney U test.

**Fig. S5:**
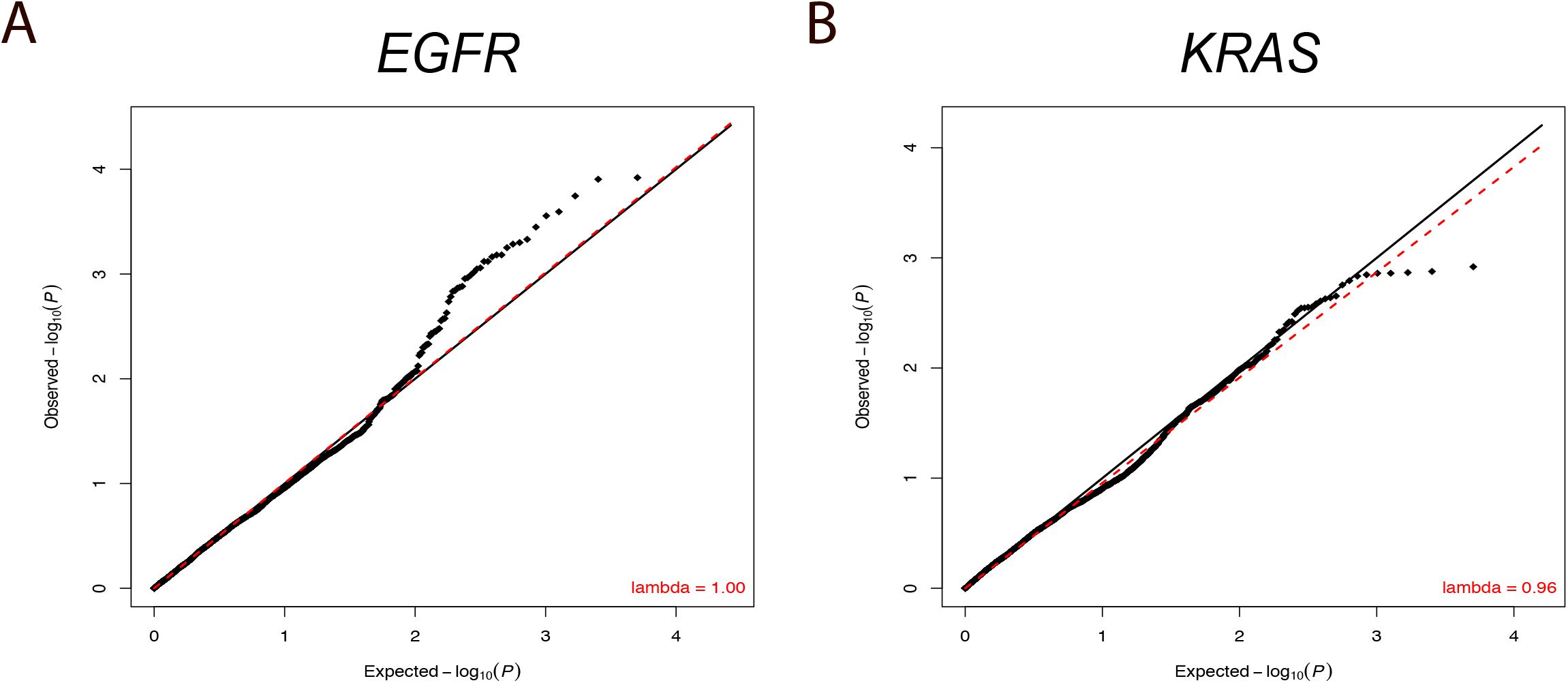
Genome-wide association of local ancestry with somatic mutations. A) QQ plots demonstrating the distribution of p values of association with *EGFR*, and B) *KRAS*.

**Fig. S6:**
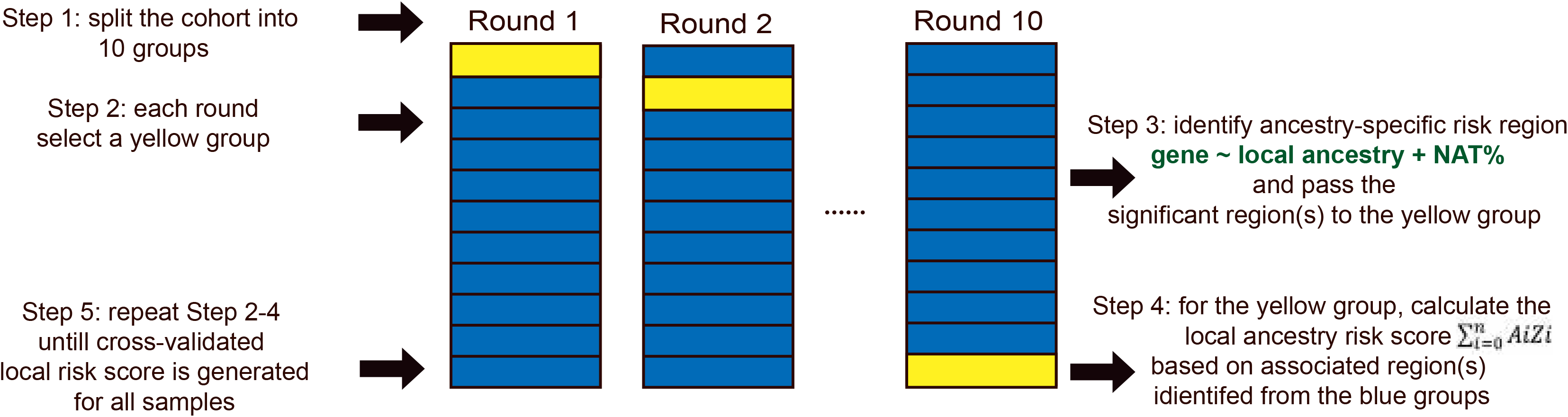
Schematic diagram for calculations of local ancestry risk scores (weighted sum of the NAT ancestry in associated genomic loci)

